# The impact of spinal surgery on the locomotive syndrome in patients with lumbar spinal stenosis in CDL stage 3: a retrospective study

**DOI:** 10.1101/2023.02.12.23285809

**Authors:** Ichiro Nakae, Ryuki Hashida, Ryota Otsubo, Sohei Iwanaga, Hiroo Matsuse, Kimiaki Yokosuka, Tatsuhiro Yoshida, Takuma Fudo, Shinji Morito, Takahiro Shimazaki, Kei Yamada, Kimiaki Sato, Naoto Shiba, Koji Hiraoka

**Affiliations:** Department of Orthopaedics, Kurume University, 67 Asahi-machi, Kurume, Fukuoka, 830-0011, Japan; Division of Rehabilitation, Kurume University Hospital, 67 Asahi-machi, Kurume, Fukuoka, 830-0011, Japan; Biostatistics Center, Kurume University, 67 Asahi-machi, Kurume, Fukuoka, 830-0011, Japan; Department of Nursing Kurume University Hospital, 67 Asahi-machi, Kurume, Fukuoka, 830-0011, Japan

**Keywords:** lumbar spinal canal stenosis, locomotive syndrome, low back pain, visual analog scale

## Abstract

**Objectives:** Locomotive syndrome (LS) is characterized by reduced mobility. Clinical decision limit (CDL) stage 3 in LS indicates physical frailty. Lumbar spinal canal stenosis (LSS) is one cause of LS, for which lumbar surgery is considered to improve the CDL stage. This study aimed to investigate the efficacy of lumbarsurgery and independent factors for improving CDL stage in patients with LSS.

**Design:** A retrospective study

**Setting:** The study was conducted at the Department of Orthopaedic Surgery at University Hospital.

**Participants:** A total of 157 patients aged ≥ 65 years with LSS underwent lumbar surgery.

**Interventions:** The 25-Question Geriatric Locomotive Function scale (GLFS-25) was used to test for LS, and the timed up and go test (TUG) was used to evaluate functional ability. Lower limb pain was evaluated using a visual analog scale. Patients with at least one improvement in CDL stage following lumbarsurgery were included in the improvement group. Differences in lower-limb pain intensity between the groups were evaluated using the Wilcoxon rank-sum test.

The Spearman’s rank correlation coefficient was used to determine correlations between Δ lower limb pain and Δ GLFS-25. Logistic regression analysis was used to identify factors associated with improvement in LS.

**Results:** The proportion of patients with improved CDL stage was 45.1% (improvement/non-improvement: 32/39). Δ Lower limb pain was significantly reduced in the improvement group compared to that in the non-improvement group (51.0 [36.3-71.0] vs 40.0 [4.0-53.5]; p =0.0107). Δ GLFS-25 significantly correlated with Δ lower-limb pain (r =0.3774, p =0.0031). In a multiple logistic regression analysis, TUG and age were significantly associated with improvement in LS (odds ratio, 1.22; 95% confidence interval: 1.07-1.47).

**Conclusions:** The lumbar surgery effectively improved the CDL stage in patients with LSS. In addition, TUG was an independent factor associated with improvement in the CDL.

**Strengths and limitations of the study:**

- The Japanese Orthopaedic Association defines locomotive syndrome as patients with reduced motor function. We reported the effect of surgical treatment on clinical decision limit 3 (CDL3), which corresponds to physical frailty.
- We investigated improvement factors for locomotive syndrome CDL stage 3 in patients with lumbar spinal stenosis in a retrospective study.
- Seventy-one patients who underwent surgical treatment were included in the study.
- Factors associated with improvement in locomotive syndrome were analyzed using multivariate logistic analysis and decision tree analysis.

## 1. INTRODUCTION

The number of older adults who require nursing care is increasing in Japan, along with the aging of a large proportion of the population. Musculoskeletal disorders resulting from fractures and falls and joint diseases are the most common causes of nursing care and can result in lost mobility. [1] In 2007, the Japanese Orthopaedic Association (JOA) defined locomotive syndrome (LS) as a condition in which mobility is reduced due to musculoskeletal disorders. [2] Later, in 2015, they established a scale to assess the severity of LS called clinical decision limit (CDL), which included stages 1 and [2] CDL stage 1 is defined as a state in which a decline in mobility has begun but may be improved through exercise habits and dietary correction. In contrast, patients in CDL stage 2 show a progressive decline in mobility. However, this classification was met with some uncertainty, as disease statuses encompassed by CDL stage 2 were broad, ranging from mild to severe. Therefore, JOA defined the additional CDL stage 3 in 2020 for early detection and therapeutic intervention in severe LS. [3] Patients in CDL stage 3 experience limited social participation owing to a significant loss of mobility. As some patients experience additional complications from orthopedic diseases and require surgery, and with the establishment of CDL stage 3, it is necessary to evaluate the extent to which LS may be improved with surgery.

The relationship between lumbar spinal canal stenosis (LSS) and LS has recently attracted much attention in spinal diseases. [4] LSS is a disease in which nerves are compressed due to intervertebral discs and joint degeneration, and thickening of the ligamentous tissue. In one Japanese cohort study, the prevalence of LSS was reported to be very high, affecting approximately 10% of adults. [5] Patients with LSS have reduced lower extremity function due to pain, numbness, and intermittent claudication, [6] which increases the risk of falling [7] and significantly impacts daily life. Lumbar spine surgeries such as decompression and spinal fusion can relieve symptoms and improve activities of daily living (ADL) in patients with LSS.

The timed up and go test (TUG) is an objective measure of functional disability that can be used to evaluate various activities such as standing, accelerating, walking, decelerating, and turning, which are often limited in patients with lumbar degenerative diseases. [8] TUG can be easily conducted with a chair and a 3-meter walking space and does not require special equipment. [9] A previous study used TUG to measure motor impairment in patients with lumbar degenerative diseases, with <11.5 seconds classified as no impairment, 11.5 to 13.4 seconds as mild impairment, 13.4 to 18.4 seconds as moderate impairment, and >18.4 seconds as severe impairment. [8] TUG is a test that is not easily affected by the patient’s mental state, lifestyle, or physique [10, 11]] and is highly related to daily life functions such as lower limb muscle strength, sense of balance, walking ability, and ease of falling. Furthermore, TUG is used to evaluate motor function in a wide range of subjects, from healthy patients to those with lumbar degenerative diseases. [11, 12] Thus, the TUG test is useful for evaluating preoperative physical function in patients with LS.

It has been reported that surgical treatment of knee joint [13] and hip joint disease [3] can improve the condition of patients in CDL Stage 3, but there have been few reports on lumbar degenerative diseases. Therefore, this study aimed to investigate the efficacy of lumbar spinal surgery for patients with LSS in CDL stage 3 and the preoperative factors associated with the improvement of the CDL stage.

## 2. MATERIALS AND METHODS

### 2.1 Patient and Public Involvement

This retrospective study was conducted between May 2020 and April 2021 at the Department of Orthopaedic Surgery at University Hospital, with 157 patients aged ≥

65 years with lumbar spinal stenosis who underwent lumbar spinal surgery without serious complications (such as ischemic heart disease or stroke). Inclusion and exclusion criteria are shown in Figure 1. Among the 157 patients involved in this study, patients in CDL stages 0, 1, and 2 were excluded (n=56). Patients with missing data were excluded (n=30). Finally, patients in CDL stage 3 were included in this study (n=71).

**Figure 1.**
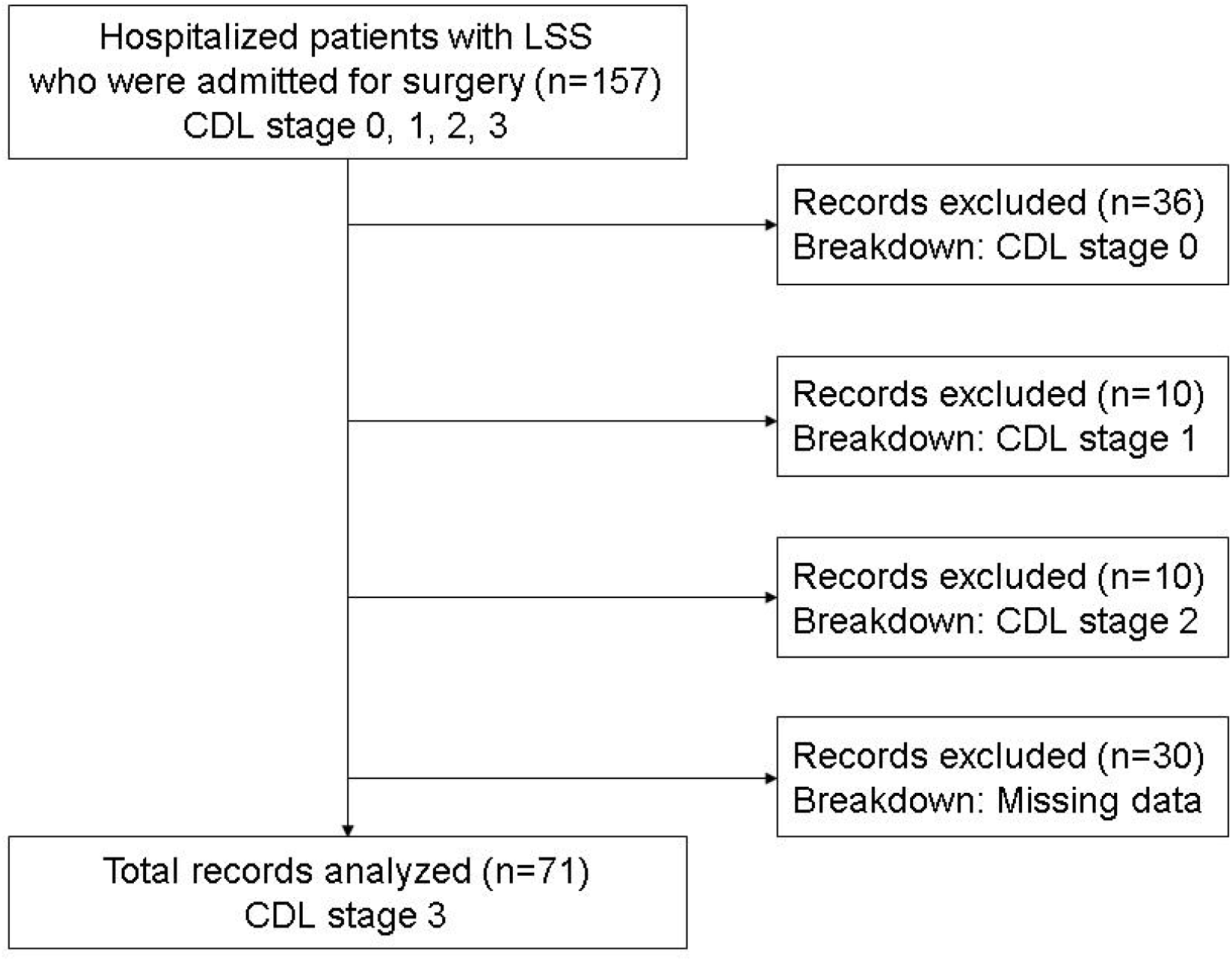
Diagram of the inclusion and exclusion criteria in this study. Abbreviations: CDL, clinical decision limit; LSS, lumbar spinal canal stenosis

This study was conducted following the Declaration of Helsinki and was approved by the Ethics Committee of the University. Consent to participate in the study was obtained using an opt-out approach.

### 2.2 Recorded data

A physical therapist performed physical function tests, and the patients completed the questionnaire. Age, sex, BMI, TUG, trunk and limb skeletal muscle mass, grip strength, life-space assessment (LSA), prognostic nutritional index (PNI), low back pain, lower limb pain, and lower limb numbness were adopted as survey items for preoperative factors related to the improvement of the LS. Furthermore, the 25-Question Geriatric Locomotive Function scale (GLFS-25) and JOA Back Pain Evaluation Questionnaire (JOABPEQ) were used to evaluate the patient before and after lumbar spinal surgery.

### 2.3 Timed Up and Go test (TUG)

TUG was conducted using a stopwatch to measure the time elapsed from when the subject’s body moved to the time they turned at a cone 3 m away and returned to sitting on the chair. A chair with a height of 42.0 cm and elbow rest were used. The subject was given the following instruction: “please get up from the chair, walk as fast as possible, turn at the cone 3 m ahead, and sit on the chair again. You may turn in either direction.”

### 2.4 Trunk and limb skeletal muscle mass

Skeletal muscle mass was assessed using the bioelectrical impedance method (Inbody720, Inbody Co., Ltd., Seoul, Korea). All patients were assessed on the day of admission. The participants grasped the handle of the analyzer with embedded electrodes and stood on a platform with the soles of their feet in contact with the electrodes (two electrodes each were placed on the feet and hands).

### 2.5 Hand grip strength

Grip strength was measured using a Smedley-type grip strength meter (Takei model T.K.K. 5101, Takei Kiki Kogyo, Akiha-ku Niigata, Japan). After hearing the signal, the participants were instructed to grip the strength meter as firmly as possible. They were instructed to “please hold the grip strength meter for 3 s.” The right side was measured twice, and the maximum value was adopted.

### 2.6 Life-Space Assessment (LSA)

The LSA was originally developed as a simple questionnaire to assess physical activity in the older adults and is a clinically useful index that has been utilized in clinical practice and research. [14] A total score is 120, with higher scores indicating higher activity levels.

### 2.7 Prognostic nutritional index (PNI)

The PNI scale reflects a patient’s inflammatory and nutritional status and is associated with postoperative complications in spinal disease. [15, 16]

### 2.8 Evaluation of pain

Low back pain, lower limb pain, and lower limb numbness were evaluated using a visual analog scale (VAS), with a score of 100 indicating extreme pain and 0 indicating no pain.

### 2.9 GLFS-25

The GLFS-25 test focuses on physical pain and ADL in the preceding month. For this study, each item was scored from “no disability” (0 points) to “severe disability” (4 points), and the total score was used to evaluate the CDL stage of the LS patient. Scores were classified as follows: stage 1, ≥ 7 points to <16 points; stage 2, ≧16 points to <24 points; and stage 3, ≧24 points. [17]

### 2.10 JOABPEQ

The Japan Orthopedics Association developed the JOABPEQ, a QOL assessment specific to lumbar spine disease that uses patient-oriented and self-reported functional status. The JOABPEQ is a disease-specific tool consisting of 25 items corresponding to five subscales: low back pain, lumbar spine function, walking ability, social function, and mental health. The score for each subscale ranges from 0-100, with higher scores indicating better conditions. [18]

### 2.11 Statistical analysis

All statistical analyses were performed using JMP version 15.0 statistical software (SAS Institute Inc., Cary, NC, USA). The GLFS-25 was used to evaluate the CDL stage. Patients with preoperative CDL stage 3 were included in the study, and those with at least one improvement in the CDL stage after lumbar spinal surgery were included in the improvement group. [19]

The Wilcoxon rank-sum test was used to compare the improved and non-improved groups. Logistic regression and decision tree analyses were used to investigate the preoperative factors associated with improvement in LS. The Spearman rank correlation coefficient was used to determine correlations between Δ lower limb pain, Δ lower limb numbness, Δ low back pain, and Δ GLFS-25. In all cases, statistical significance was set at p <0.05. All data are expressed as the median (interquartile range) and range.

## 3. RESULTS

### Characteristics of Patients with LSS

Of the 157 participants who met the eligibility criteria for this study, 71 patients’ data were analyzed after applying the exclusion criteria. Patient characteristics are shown in Table 1.

**Table 1:** The characteristic of the patients.

|  | Clinical Decision Limits stge3<br>(N=71) |  |
| --- | --- | --- |
|  | Median | Range (min-mx) |
| Age (years) | 77 | 65-91 |
| Sex (Male/Female) | 27/44 |  |
| BMI (kg/m<sup>2</sup>) | 23.6 | 15.4-32.1 |
| Limb skeletal muscle mass (kg) | 13.9 | 8.9-23.3 |
| Trunk skeletal muscle mass (kg) | 16.6 | 11.7-23.7 |
| Hand grip strength (kg) | 21.4 | 8.1-44 |
| TUG (s) | 11.5 | 5.4-38.4 |
| LSA | 48 | 0-120 |
| PNI | 51.9 | 36.2-64.9 |
| Low back pain (VAS) | 63 | 0-100 |
| Lower limb pain (VAS) | 68 | 0-100 |
| Lower limb numbness | 60 | 0-100 |
| (VAS) |  |  |
| GLFS-25 | 44 | 24-87 |

Abbreviation: BMI; Body Mass Index, TUG; Timed up and go test,
LSA; Life space assessment, PNI; Prognostic nutritional index
VAS; Visual Analog Scale, GLFS-25; the 25-Question Geriatric Locomotive Function Scale
\*: means statistically significant

### Surgical Results of patients with LSS

Figure 2 shows the breakdown of preoperative and postoperative GLFS-25 scores and CDL stages at three months postoperatively. Lumbar spinal surgery improved the mean GFLS-25 score from 48.8 preoperatively to 30 postoperatively (Fig. 2A). The rates of CDL stages at three months postoperatively were as follows: CDL stages 0, 1, 2, and 3 were 7%,18.3%, 19.7%, and 55%, respectively. In total, the proportion of patients with improved CDL stage was 45.1% (32/71) (Fig. 2B).

**Fig. 2-A.**
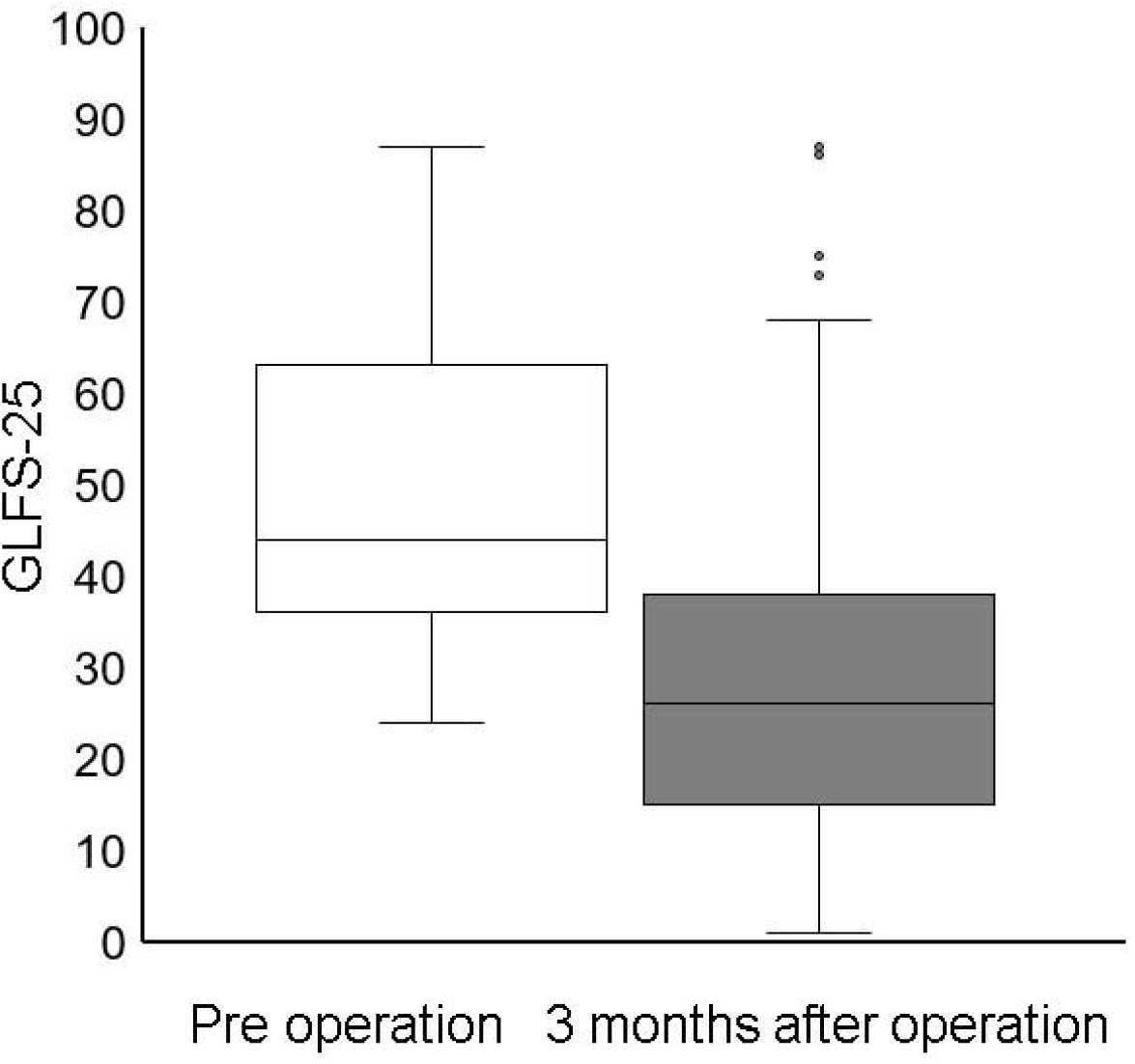
The mean GLFS-25 scores preoperatively and 3 months postoperatively.

**Fig. 2-B.**
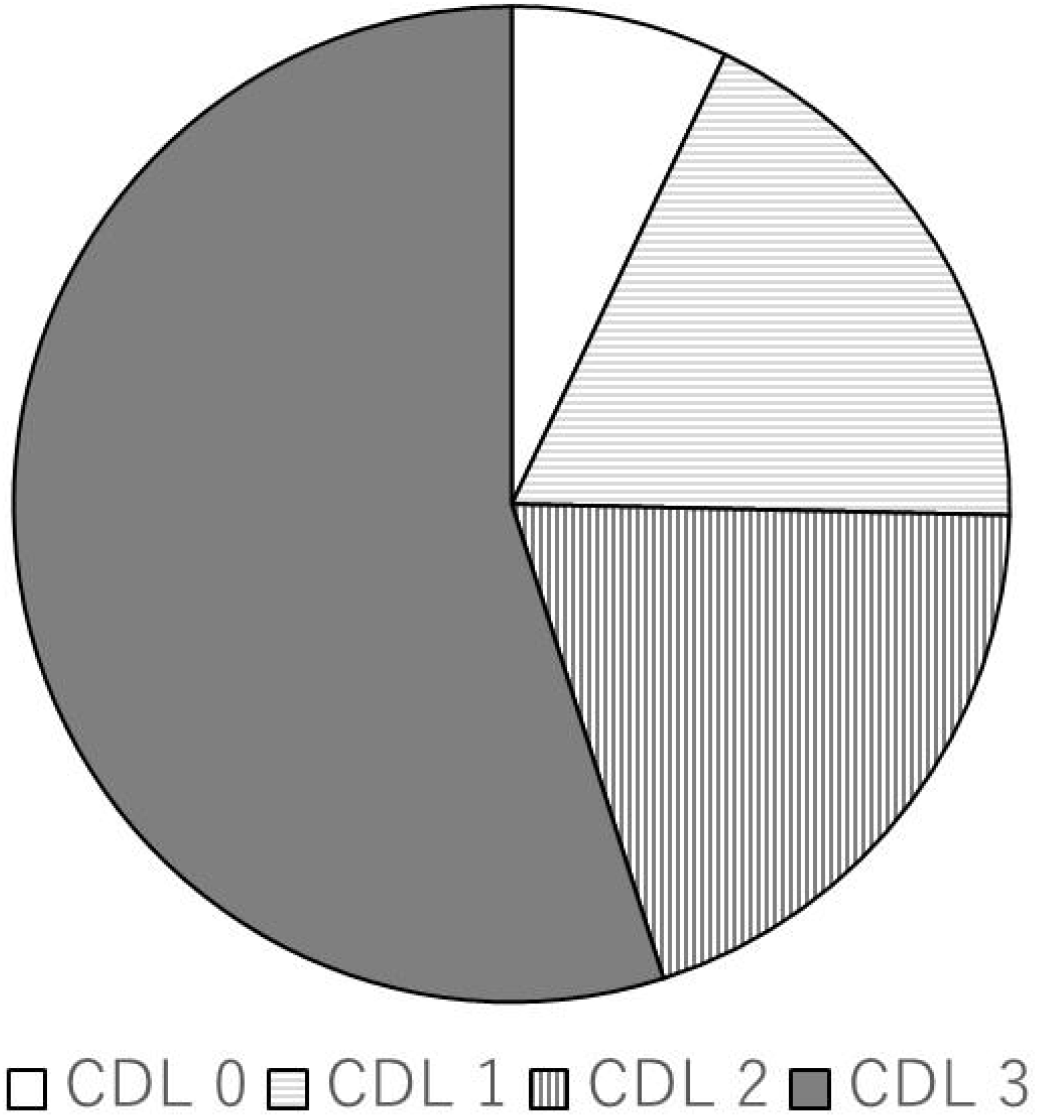
The ratio in patients’ CDL stage at 3 months postoperatively. Abbreviations: CDL, clinical decision limit; GLFS-25, 25-Question Geriatric Locomotive Function Scale

### Comparison of preoperative status between two groups

Patients in the improvement group were significantly younger than those in the non-improvement group. In addition, the LSA score was significantly higher in the improvement group than in the non-improvement group (Table 2). The TUG times in the improvement group were significantly shorter than that in the non-improvement group. The two groups had no significant differences in limb skeletal muscle, trunk skeletal muscle, PNI, or VAS scores.

**Table 2:** Comparison between two groups.

|  | Improvement group (N=32) |  | Non-improvement group (N=39) |  | P |
| --- | --- | --- | --- | --- | --- |
|  | Median | Range (min-max) | Median | Range (min-max) |  |
| Age | 75 | 65-81 | 78 | 67-91 | 0.008 |
| Sex (Male/Female) | 14/18 |  | 13/26 |  | 0.4 |
| BMI (kg/m<sup>2</sup>) | 22.8 | 15.4-32.1 | 24.2 | 17.7-31.2 | 0.3 |
| Limb skeletal | 14.9 | 9.8-23.3 | 13.9 | 8.9-21.9 | 0.3 |
| muscle mass (kg) |  |  |  |  |  |
| Trunk skeletal muscle mass (kg) | 16.8 | 12.4-23.3 | 16.5 | 11.7-23.7 | 0.3 |
| Hand grip strength (kg) | 23.3 | 12.8-44 | 20.6 | 8.1-43 | 0.1 |
| TUG (s) | 9.6 | 5.4-21.5 | 14.5 | 6.4-38.4 | 0.0002 |
| LSA | 54 | 4.5-120 | 37.5 | 0-110 | 0.02 |
| PNI | 52.4 | 36.9-64.9 | 51.2 | 36.2-60.5 | 0.2 |
| Low back pain (VAS) | 51 | 8-100 | 67 | 0-100 | 0.2 |
| Lower limb pain (VAS) | 65 | 18-100 | 69.5 | 0-100 | 0.8 |
| Lower limb numbness (VAS) | 51 | 0-100 | 65.5 | 0-100 | 0.4 |
| GLFS-25 | 40 | 24-65 | 58 | 25-87 | 0.002 |

Abbreviation: BMI; Body Mass Index, TUG; Timed up and go test, LSA; Life space assessment, PNI; Prognostic nutritional index, VAS; Visual Analog Scale, GLFS-25; the 25-Question Geriatric Locomotive Function Scale
\*: means statistically significant

With the exception of pain and lumbar function in the non-improvement group, lumbar spinal surgery improved the JOABPEQ scores in both groups. Similarly, lumbar spinal surgery improved the VAS scores for low back pain, lower limb pain, and lower limb numbness in both groups (Supplementary table).

### Cox regression analysis for improvements in LS

Multivariate regression analysis was performed with the following variables related to improvements: age, sex, BMI, LSA, PNI, handgrip strength, and TUG.

The TUG and age were significantly associated with improvement in LS (p =0.0017 and p =0.031) (Table 3). LSA, BMI, PNI, and handgrip strength were not significantly associated with improvements in LS (p =0.38, p =0.45, p =0.61, p =0.65).

**Table 3:** Multivariate analysis for Improvement of locomotive syndrome.

| Factors | Multivariate analysis for<br>Improvement of locomotive<br>syndrome<br>(95% Confidence interval,<br>P-value) |
| --- | --- |
| TUG | 1.22<br>(1.07-1.47, 0.0017) |
| Age | 1.15<br>(1.01-1.32, 0.031) |
| LSA | 1.01<br>(0.99-1.04, 0.38) |
| BMI | 1.07<br>(0.90-1.28, 0.45) |
| PNI | 1.04<br>(0.90-1.21, 0.61) |
| Hand Grip strength | 1.02<br>(0.94-1.12, 0.65) |

Abbreviation: TUG; Timed up and go test, LSA; Life space assessment, BMI; Body Mass Index, PNI; Prognostic nutritional index,
\*: means statistically significant

### Decision-tree algorithm for improvement of LS

The decision-tree algorithm was performed with variables related to improvements, including age, sex, BMI, LSA, PNI, handgrip strength, and TUG. The TUG test was selected as the first divergence variable in the decision tree analysis and LSA as the second divergence variable. Among patients with TUG < 12.4, CDL stage 3 was improved in 66.7%, and LSA was the second divergence variable in patients with TUG ≥ 12.4. Patients with TUG ≧ 12.4 who had LSA ≥ 40 showed improvement from CDL stage 3 by 46.2%. Among patients with reduced physical function, CDL stage 3 was improved upon in 46.2%, with an LSA score of 40 or higher (Fig. 3).

**Fig. 3:**
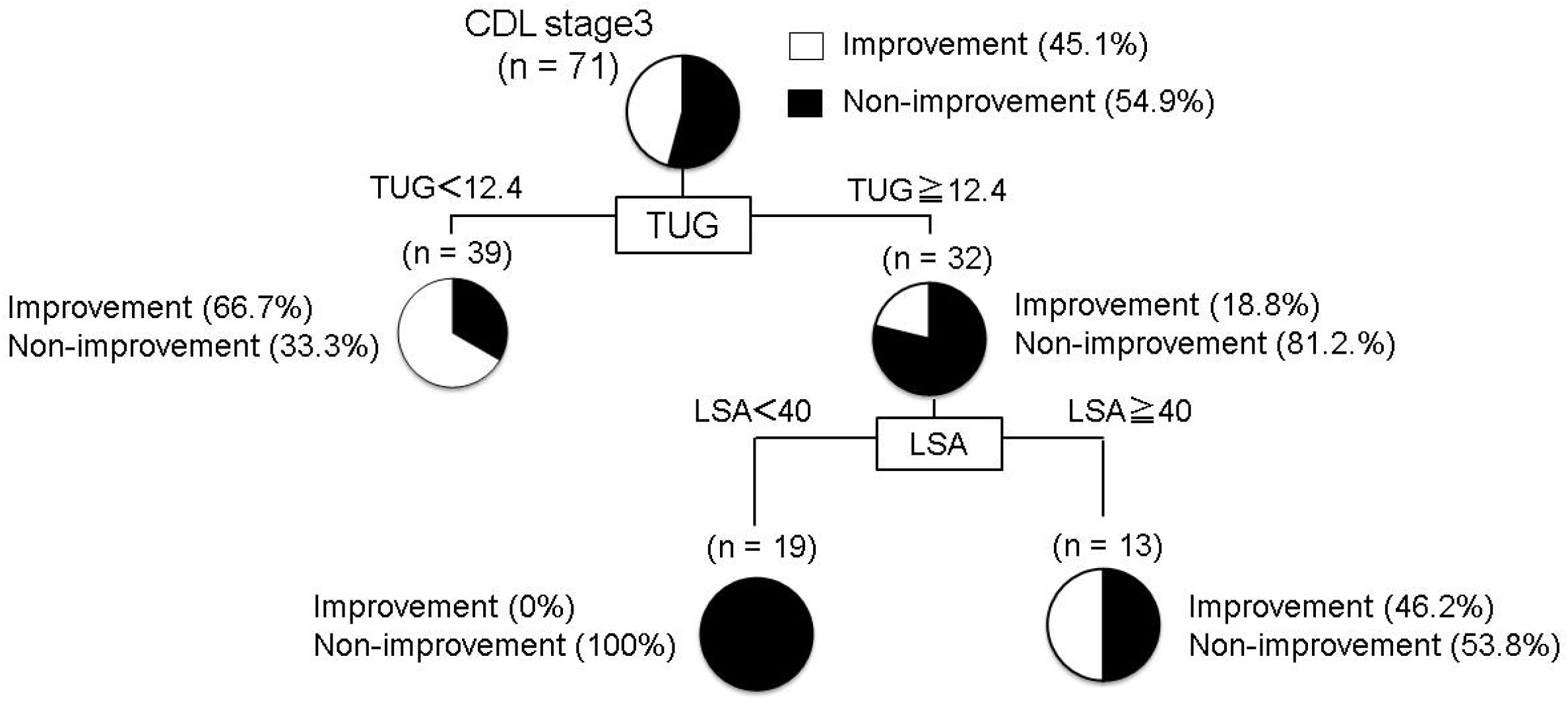
Decision tree analysis for preoperative factors associated with improvement in locomotive syndrome. Abbreviations: CDL, clinical decision limit; LSA, life space assessment; TUG, timed up and go test

### Relationship between breakdown of CDL stage and preoperative TUG three months postoperatively

Figure 4 shows the relationship between the breakdown of the CDL stage and preoperative TUG 3 months following lumbar spinal surgery. In patients with CDL stage 3 LS, the preoperative TUG time 3 months following the procedure was significantly longer than that for those in CDL stages 0 and 1 (Fig. 4). The results showed that patients who had low preoperative TUG scores improved to either CLD stage 0 or 1 from stage 3 following the surgical procedure.

**Fig. 4:**
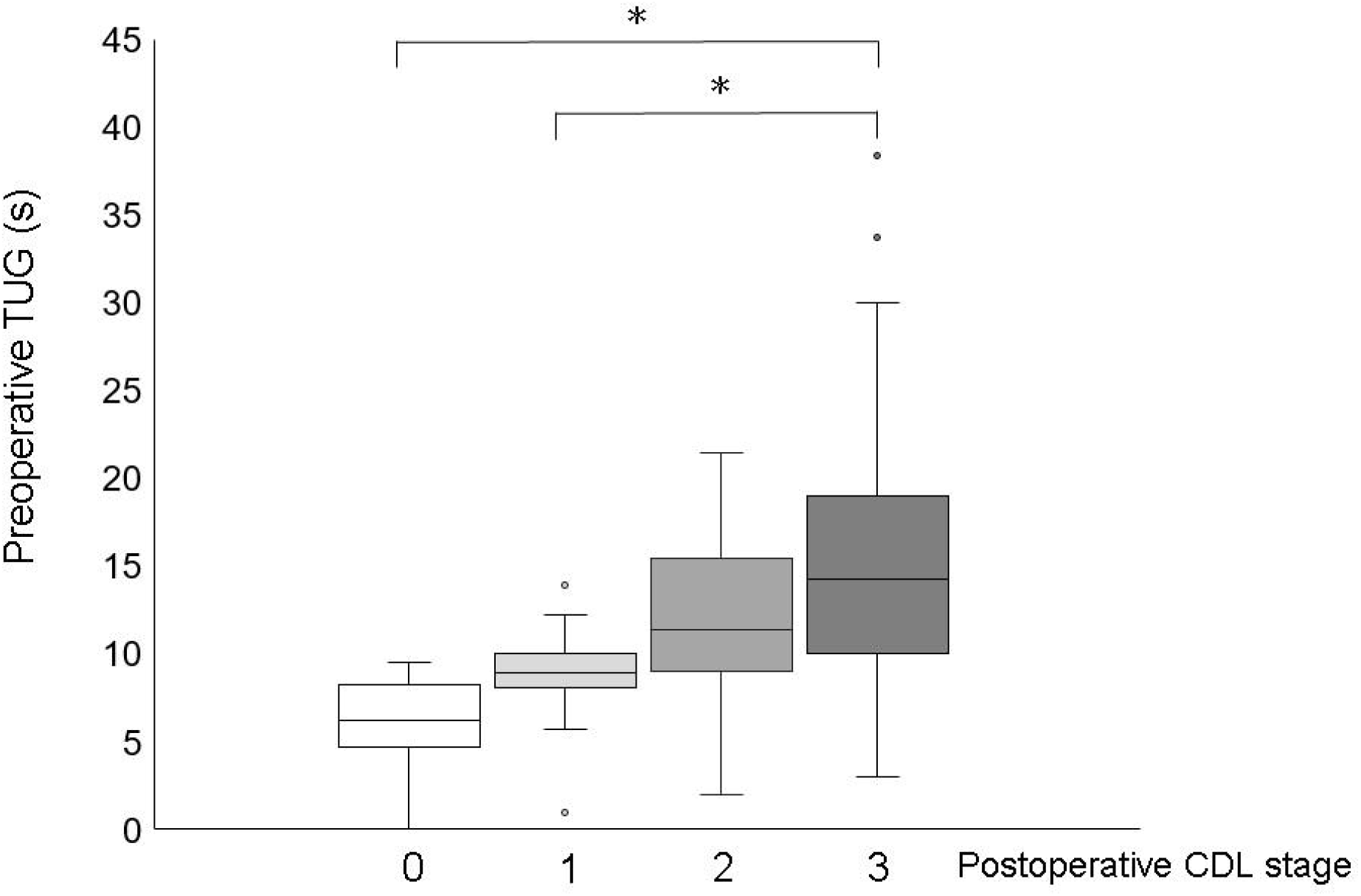
The relationship between the breakdown of the CDL stage and preoperative TUG at three months after lumbar spinal surgery. Abbreviations: CDL, clinical decision limit; TUG, timed up and go test *: means statistically significant

### Relationship between Δ GFLS-25 and Δ pain scales

Δ GLFS-25 was significantly positively correlated with Δ lower limb pain, Δ lower limb numbness, and Δ low back pain (Table 4).The numbers in the table indicate correlation coeffect

**Table 4:** The relationship between the delta 25-Question Geriatric Locomotive Function Scale and each delta pain scale.

| | $\Delta$ Lower limb pain | $\Delta$ Lower limb numbness | $\Delta$ Low back pain | $\Delta$ GLFS-25 |
| --- | --- | --- | --- | --- |
| $\Delta$ Lower limb pain | 1.0000 | | | |
| $\Delta$ Lower limb numbness | 0.5684* | 1.0000 | | |
| $\Delta$ Low back pain | 0.4479* | 0.3315* | 1.0000 | |
| $\Delta$ GLFS-25 | 0.3615* | 0.3723* | 0.4103* | 1.0000 |

Abbreviation: GLFS-25; the 25-Question Geriatric Locomotive Function Scale

### Comparison of Δ GLFS-25 and Δ pain scales between groups

Lower limb pain and GFLS-25 scores were significantly better in the improvement group than in the non-improvement group (p =0.0107, p =0.002). Low back pain and lower limb numbness in the improvement group were not significantly higher than those in the non-improvement group (p =0.0953, p =0,1041) (Fig. 5).

**Fig. 5:**
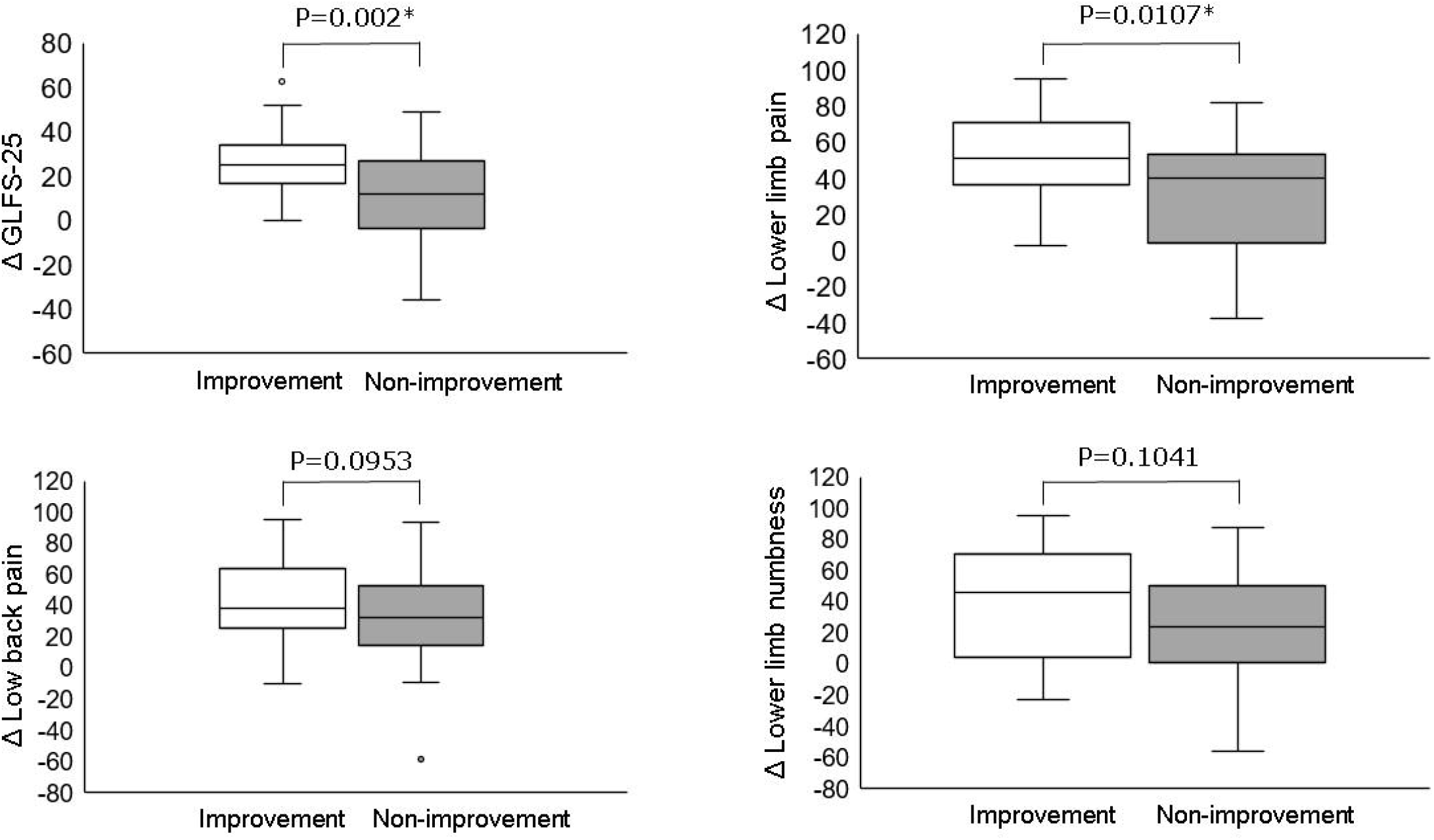
Comparison between the two groups on Δ GLFS-25, Δ Lower limbs pain, Δ Low back pain, and Δ Lower limb numbness. Abbreviations: GLFS-25; 25-Question Geriatric Locomotive Function Scale *: means statistically significant

## DISCUSSION

In this study, the rate of CDL stage improvement achieved by lumbar spinal surgery was 45.1% (32/71) in patients with CDL stage 3. The VAS score for lower extremity pain was significantly improved in the group with an improved CDL stage. Moreover, the Δ GLFS-25 was significantly correlated with the Δ VAS of all pain scales. Thus, the improvements to LS might have been associated with a reduction in pain. The reduction in pain achieved by lumbar spinal surgery improved the ADL and CDL stages. Furthermore, the preoperative TUG test was the most relevant factor for the postoperative results. Preoperative evaluation of physical function is useful for determining surgical indications.

The LS is caused by various orthopedic diseases, resulting in reduced quality of life and shortened expectancy of healthy life. [2] Fujita et al. reported that LSS is a potential risk factor for LS [20] and that lumbar spinal surgery effectively reduces the LS risk. [21] The change in the GLFS25 score in this study was comparable to that reported in previous studies. [21-23] However, in this study, lumbar spinal surgery significantly improved not only the GLFS25 score, but also the VAS score for low back pain and lower limb pain (Supplementary data). Therefore, lumbar spinal surgery may ameliorate LS by improving pain and numbness and enhancing the quality of life in patients with LSS. Moreover, the improvement rate of CDL stage via lumbar spinal surgery in this study was 45.1% (32/71). Among the patients who received the surgery, 7% (5/71) had outgrowth LS. Additionally, it was found that 7% of the patients who needed care due to lumbar spinal surgery no longer had LS.

Moderate physical activity is important for maintaining and improving life functions and preventing disease progression, disability, and frailty In older adults. [24-26] In addition, previous studies have reported that LSA is significantly correlated with ADL in community-dwelling older individuals. [14, 27] A cohort study of relatively healthy community-dwelling older people reported LSA scores of 62.9±24.7 [14] or 64.1±24.7. [27] Therefore, it can be inferred that patients with LSS in this study were more physically inactive as they had lower LSA scores (median score of 48) than those in previous studies. In the decision tree analysis, even in patients with reduced physical function (TUG ≥ 12.4 seconds), CDL stage 3 was improved with a 50% probability if the LSA was 40 or higher. Therefore, increasing preoperative physical activity may have affected the postoperative outcomes.

This study aimed to identify preoperative factors associated with the improvement of CDL stage 3 in patients with LSS. In the decision tree analysis, the preoperative factor most associated with LS improvement was the TUG test score. In this study, five of 71 patients (7%) got rid of their LS 3 months after lumbar spinal surgery (Figure 2). The median TUG time of these patients was 6.2 seconds, and their preoperative physical function was clearly higher than that before the procedure (Figure 4). In this study, the Δ VAS score for low back pain in the improvement group tended to be higher than that in the non-improvement group. Moreover, the Δ VAS score for lower-limb pain in the improvement group was significantly higher than that in the non-improvement group. The Δ VAS score for low back and lower limb pain was correlated with the Δ GLFS-25 score (Table 4), and the total GLFS-25 score was associated with pain in the patients with LSS. [28] Thus, improving low back and lower limb pain through lumbar spinal surgery might improve GLFS-25 scores. These patients may have been identified as being in CDL stage 3 because their ADL was severely limited due to pain, although their physical function was not impaired. Improved low back and lower limb pain due to lumbar spinal surgery may have dramatically improved ADL and allowed patients to break free from LS. However, the results of the non-improvement group suggested that it was challenging to improve LS by lumbar spinal surgery in patients who had experienced a significant preoperative decline in physical function prior to the surgery. In summary, older patients with CDL stage 3 LSS without any preoperative decline in physical function may have a good postoperative course in which they no longer have LS; such patients should be encouraged to undergo lumbar spinal surgery earlier. It has been suggested that preoperative physical function assessment is important in determining the indication for lumbar spinal surgery and that TUG is an important factor among them.

This study had some limitations. First, it was a single-center study. Second, owing to the small sample size, we were unable to divide the patients into decompression and lumbar fusion groups. Third, we assessed LS using only the GLFS-25; however, this is difficult to assess by 2 step and stand-up tests in patients with the severe LS. Kato et al. showed the use of the GLFS-25 assessment to be appropriate for patients with several LS and musculoskeletal diseases requiring surgery. [29]

This study showed that improvement in the low back and lower limb pain through lumbar spinal surgery might be beneficial for improving CDL stage 3. TUG score was an independent factor associated with the improvement of CDL stage 3 in patients with lumbar spinal canal stenosis.

## Data Availability

Hashida, Ryuki, 2022, The impact of spinal surgery on the locomotive syndrome in patients with lumbar spinal stenosis, https://doi.org/10.7910/DVN/QPA7XO, Harvard Dataverse, DRAFT VERSION

## Acknowledgments

None

## Author contributions

IN, RH, participated in study conception and design, interpretation of data, and drafting of the manuscript. SI, RO, TF, SM performed data acquisition. KY, TY, TS, KY, KH, KS, HM, NS participated in interpretation of data and critical revision.

## Reporting statement

### Patient consent form

Consent to participate in the study was obtained using an opt-out approach. No identifying information of the patients has been shared.

### Data sharing statement

Hashida, Ryuki, 2022, “The impact of spinal surgery on the locomotive syndrome in patients with lumbar spinal stenosis”, https://doi.org/10.7910/DVN/QPA7XO, Harvard Dataverse, DRAFT VERSION

### Research Ethics Approval: Human Participants

This study was conducted in accordance with the Herkinsi Declaration and was approved by the Ethics Committee of Kurume University (Approval ID:22025)

