## supplementally table for "The impact of spinal surgery on the locomotive syndrome in patients with lumbar spinal stenosis in CDL stage 3: a retrospective study"

Supplementary Table

|  | Improvement group (N=32) | | | | P | Non-improvement group (N=39) | | | | P |
| --- | --- | --- | --- | --- | --- | --- | --- | --- | --- | --- |
|  | Pre operation | | Three months  after operation | |  | Pre operation | | Three months  after operation | |  |
|  | Median | Range  (min-max) | Median | Range  (min-max) |  | Median | Range  (min-max) | Median | Range  (min-max) |  |
| JOABPEQ |  |  |  |  |  |  |  |  |  |  |
| Low back pain | 43 | 0-100 | 100 | 14-100 | 0.0002 | 43 | 0-100 | 71 | 0-100 | 0.08 |
| Lumbar function | 50 | 8-100 | 83 | 25-100 | <0.0001 | 42 | 0-100 | 42 | 0-100 | 0.9 |
| Walking ability | 29 | 0-93 | 67.5 | 21-100 | <0.0001 | 14 | 0-86 | 43 | 0-86 | 0.01 |
| Social life function | 44.5 | 3-92 | 65 | 38-100 | <0.0001 | 32 | 0-86 | 49 | 0-95 | 0.02 |
| Mental health | 43.5 | 6-76 | 58.5 | 42-90 | <0.0001 | 42 | 0-79 | 48 | 0-83 | 0.01 |
| VAS (mm) |  |  |  |  |  |  |  |  |  |  |
| Low back pain | 51 | 8-100 | 11 | 0-53 | <0.0001 | 67 | 0-100 | 25.5 | 0-81 | <0.0001 |
| Lower limb pain | 65 | 18-100 | 9 | 0-58 | <0.0001 | 69.5 | 0-100 | 30.5 | 0-87 | <0.0001 |
| Lower limb numbness | 51 | 0-100 | 10.5 | 0-44 | <0.0001 | 65.5 | 0-100 | 31.5 | 0-100 | <0.0001 |

Abbreviation:

JOABPEQ; Japanese Orthopaedic Association Back Pain Evaluation Questionnaire

VAS; Visual Analog Scale

*: means statistically significant
